# Results from Integrating Gender-Based Violence Services into HIV Care – a Case Study of Lighthouse Trust ART Clinic in Malawi

**DOI:** 10.1101/2024.04.26.24306348

**Authors:** Christine Kiruthu-Kamamia, Tapiwa Kumwenda, Joseph Lungu, Odala Sande, Joseph Diele, Ellen MacLachlan, Agnes Thawani

**Author notes:** Author Contributions Conceptualization: CKK Data Curation: OS, TK Formal Analysis: CKK, OS Investigation: OS, CKK Methodology: CKK, OS Project Administration: TK, JD, JL Supervision: AT Validation: CKK, OS Visualization: CKK Writing: Original Draft Preparation: CKK Writing Review & Editing: CKK, EM, AT. Funding statement: N/A. Competing interests: None.

## Abstract

**Introduction:** Gender-based violence (GBV) not only poses significant public health and human rights challenges but is also closely associated with HIV. GBV acts as a barrier to HIV prevention, testing, and treatment adherence, and fear of GBV inhibits disclosure of HIV status to sexual partners. In Malawi, where both GBV and HIV prevalence is high, integrating GBV services into HIV care is crucial. We describe the integration of GBV services into Lighthouse Trust’s ART clinics in Malawi, including screening, documentation, program implementation, and outcomes.

**Methods:** We conducted a retrospective analysis from January 2020 to September 2023. Data on cases identified, post-GBV services, and perpetrator demographics were collected from the GBV register. We used descriptive statistics to describe the program outcomes.

**Results:** We documented 7148 reported GBV cases from January 2020 to September 2023. Young women, particularly those aged 10-19, constituted a significant proportion of survivors. Psycho-social services were the most common type of service that was offered to GBV survivors (25%), followed by HIV testing (19%) and STI screening(18%). Perpetrators were mostly known to survivors or intimate partners of the survivor.

**Conclusion:** We successfully integrated GBV services into the Lighthouse Trust ART clinic, in close collaboration with the one-stop centers. Training healthcare providers enhanced support for GBV survivors, with a focus on increasing awareness, especially for minors. Recommended actions include improving access to services, confidentiality, and multi-sectoral collaboration to ensure comprehensive care aimed to create a safer, more dignified healthcare environment for all, particularly GBV survivors.

**Key Messages:** *Key Findings Teaser Key Message:*

- We successfully integrated GBV services into HIV care via adequate training, screening and identification procedures, and documentation.
- Integrating GBV screening into ART care identified over 7000 cases with most cases of sexual violence in women aged 10 to 19 years.
- There is still a need for public awareness of GBV and access to support services.

*Key Messages and Implications to policy, program managers, donors etc:*

- We provide a replicable model for healthcare providers and facility administrators, demonstrating the effectiveness of standardized screening, care, and referral processes in improving the support and outcomes for GBV survivors in low-resource settings.
- Our findings can inform the development of more inclusive health policies and support services that address the needs of all GBV survivors.

## Background

Gender-based violence (GBV) is defined as violence that results in, or may result, in physical, sexual or psychological harm or suffering to individuals based on their gender women^1^. GBV is not just a major public health problem but also a human rights problem. This violence is deeply rooted in structural, gender-based inequalities that are perpetuated by social and political forces.^2^ The four main types of GBV are physical, sexual, emotional or psychological, and economic, all of which have been linked to a broad range of mental and physical health problems, including HIV.^3^

In Malawi, GBV remains a pervasive issue, with all four types of GBV present in varying degrees^4^. A 2013 survey found that one out of five females and one out of seven males in Malawi have experienced at least one incident of sexual abuse before the age of 18 ^5^.

To support GBV survivors, the government of Malawi established One-Stop Centers (OSCs) surrounding select hospitals, where comprehensive post-GBV care is provided, including clinical, psycho-social, and legal support.^6^ These OSCs are detached buildings that provide space for healthcare providers, social workers, and police, all available in the same location to manage GBV cases.

In Malawi, where HIV prevalence is high^7^, the intersection of GBV with HIV/AIDS becomes particularly critical to address.

Research has demonstrated that exposure to GBV is closely associated with poor health outcomes^8,^ including HIV/AIDS acquisition and treatment interruption^9^. GBV acts as a barrier to HIV prevention, testing, and treatment adherence, and fear of GBV has been shown to inhibit disclosure of HIV status to sexual partners^10–12^. Routine screening for GBV in healthcare settings is not recommended due to insufficient evidence on improved outcomes for women, especially if there is no staff capacity or referral mechanisms available ^8,13^. However, some studies have shown that women are receptive to GBV screening and it can help identify those at risk of violence^14^ . Given that HIV increases the risk of GBV, it is essential to integrate GBV services into HIV care to address the intersecting needs of individuals affected by both HIV and GBV.

Many GBV interventions have focused on identifying and reducing partner violence, primarily in antenatal care or primary health care settings^15,16^. However, few studies have evaluated comprehensive GBV integration within HIV services across the cascade^17–20^ Integrating GBV into HIV care demands a carefully coordinated approach that caters to both the health and psychological needs of survivors^8^. However, documented best practices for such GBV integration in low-resource settings are still limited. Lessons learned from integrating GBV response into ART services can provide healthcare providers practical guidance on how to effectively incorporate GBV screenings, referrals, and care in HIV care. These insights can help shape protocols, inform training programs, and influence policy, ensuring that healthcare providers are equipped to offer comprehensive care that addresses the physical and emotional needs of all GBV survivors.

Thus, this study aims to describe GBV service integration at Lighthouse Trust ART clinic in Malawi through analysis of routinely collected data. In this paper, we describe how we integrated GBV services into Lighthouse Trust ART clinics and report on the GBV screening process, documentation, program implementation, and results. By examining this integration, we aim to contribute to the existing body of knowledge and inform future interventions and policies aimed at effectively addressing GBV within the context of HIV care.

## Methods

### Setting

The Lighthouse Trust is a World Health Organization (WHO)-recognized Center of Excellence (COE) and the largest HIV clinic in Malawi.^21–23^ The Lighthouse Trust operates four COEs: two in Lilongwe at the Lighthouse Clinic at Kamuzu Central Hospital (KCH) and, the Martin Preuss Center (MPC) at Bwaila Hospital, one in Blantyre at the Umodzi Family Clinic (UFC) at Queen Elizabeth Central Hospital (QECH), and one in Zomba at the Tisungane Clinic at Zomba Central Hospital (ZCH) and one in Mzuzu at Rainbow Clinic at Mzuzu Central Hospital^24^ . Four COEs are strategically located within tertiary central hospitals at KCH, QECH, ZCH, and MCH, where OSCs have been established. Additionally, the Lighthouse Trust supports eight MOH-operated health centers in Lilongwe, including Area 18, Chileka, Chitedze, Kawale, KCH, Lumbadzi, Mitundu, and Nathenje^24^.

### GBV screening process at Lighthouse Trust

Based on WHO guidelines^8^, Lighthouse does not conduct universal screening for GBV. At all clinic service delivery points, including the front desk and guard stations, healthcare providers utilize a standardized screening tool (refer to Appendix A: GBV Screening Tool) to assess the occurrence of GBV among patients. Additionally, providers undergo training to recognize subtle behaviors, health issues, or services sought that might indicate the presence of GBV (refer to Appendix B: Physical and emotional signs of potential GBV experience). GBV screening is only conducted when providers identify such indications. To ensure the accuracy and consistency of GBV screening procedures, Lighthouse developed a detailed standard operating procedure (SOP) (refer to Appendix C: GBV Screening SOP) that outlines the appropriate protocols for administering GBV screening.

### Referral to post-GBV care at Lighthouse Trust

Once clients are identified as having experienced GBV, they are referred to the appropriate provider for additional support. Each COE is staffed with a focal person for post-GBV care. Psychosocial care may be provided by referring the client to a designated psychosocial counselor (refer to Appendix D: Psychosocial counseling for GBV survivors SOP). In cases where the client has experienced severe physical violence, a referral is made to the central hospital for advanced medical care. Furthermore, the client may be referred to the OSC, where a police officer is available for judicial support. Before making a referral, the client is given adequate time to decide whether to accept the referral. If the client is not interested in any of the referral services, providers do not exert pressure but instead provide information about available post-GBV care and educate the client on the mental and physical health effects of GBV. Providers offer to schedule a follow-up visit if the client is willing. Lighthouse provides the following post-GBV clinical care to all clients that screen positive for GBV: HIV testing services (HTS), Post-exposure prophylaxis (PEP), sexually transmitted infections (STI) screening and treatment, psycho-social services (PSS) and emergency contraception (EC).

## The One-Stop-Center

The OSC is a purposefully designed facility situated within a hospital setting, with the primary objective of providing comprehensive, multidisciplinary services to survivors of GBV. These services include prompt medical and mental healthcare, immediate law enforcement assistance, and social welfare agencies’ support. Evidence has shown that establishing such centers is significantly associated with heightened prosecution rates^25^. In Malawi, the OSC caters to both adult and child survivors, addressing various forms of abuse. To ensure the delivery of comprehensive care, OSC operations necessitate effective coordination and communication among a multidisciplinary team. This team comprises professionals from diverse fields, including medical care, social welfare, mental health, and the judiciary, who collaborate to address the multifaceted needs of survivors.

In instances where specific roles within the team remain unfilled, such as the absence of a police unit in a rural catchment area, a referral protocol is used to facilitate clear communication and delineate referral pathways, thereby ensuring the availability of requisite services for survivors, irrespective of geographical constraints.

## Training

Three distinct training programs were implemented to enhance the capabilities of key healthcare providers, including nurses, clinicians, HIV diagnostic assistants, and psycho-social counselors. The first training focused on GBV and the OSC function, conducted by facilitators from the Ministry of Gender, the Ministry of Health, the Judiciary, and the Police. The second training program, conducted by the Police-Victim Support Unit, addressed GBV and child protection. Lastly, the third training was on use of the LIVES (Listen, Inquire, Validate, Enhance safety, and Support)^17^ model which two LH GBV Team Leads conducted. The LIVES training, an initiative by the WHO, aimed to assist providers in identifying instances of violence within a clinical setting and providing initial support. These comprehensive training sessions sought to equip healthcare providers with a holistic understanding of survivor management. The trainings included various topics, such as describing different forms of GBV, GBV screening using a tool adapted from Papua New Guinea^26^ Guyana, offering initial support, guidelines for reporting incidents to the police, child rights, comprehensive healthcare packages, forensic examinations, courtroom testimony, and proper documentation in the GBV register. In total, 174 providers were trained.

## Data collection and analysis

The OSCs did not possess an official register to record GBV cases prior to the start of this project . Instead, cases were documented in a hardcover notebook that was utilized for several years, resulting in a nearly torn cover. Furthermore, gaps in available data and information indicated the need to enhance GBV documentation processes. To address this issue, Lighthouse developed a comprehensive and user-friendly GBV register incorporating essential indicators for both the OSC and Lighthouse Trust (see Figure 1).

**Figure 1:**
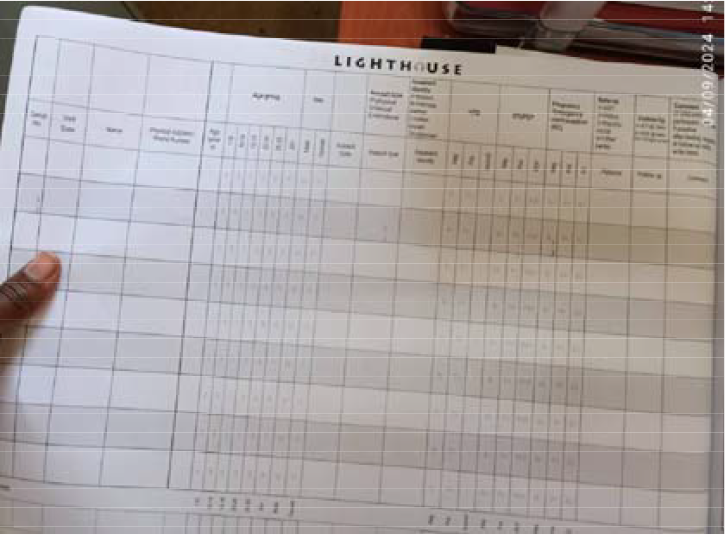
GBV Register. Data was collected from the Lighthouse Trust GBV register from January 2020 – September 2023. A retrospective analysis was conducted using descriptive statistics to analyze the data using Microsoft Excel.

## Results

From January 2020 to September 2023, Lighthouse COEs (LH, MPC, UFC, Tisungane clinic) and supported facilities (Kawale and KCH) reported 7148 cases of GBV. Most of the GBV cases were reported among the 10-14 and 15-19 age groups (see Figure 2). Approximately 93% of survivors were female.

**Figure 2:**
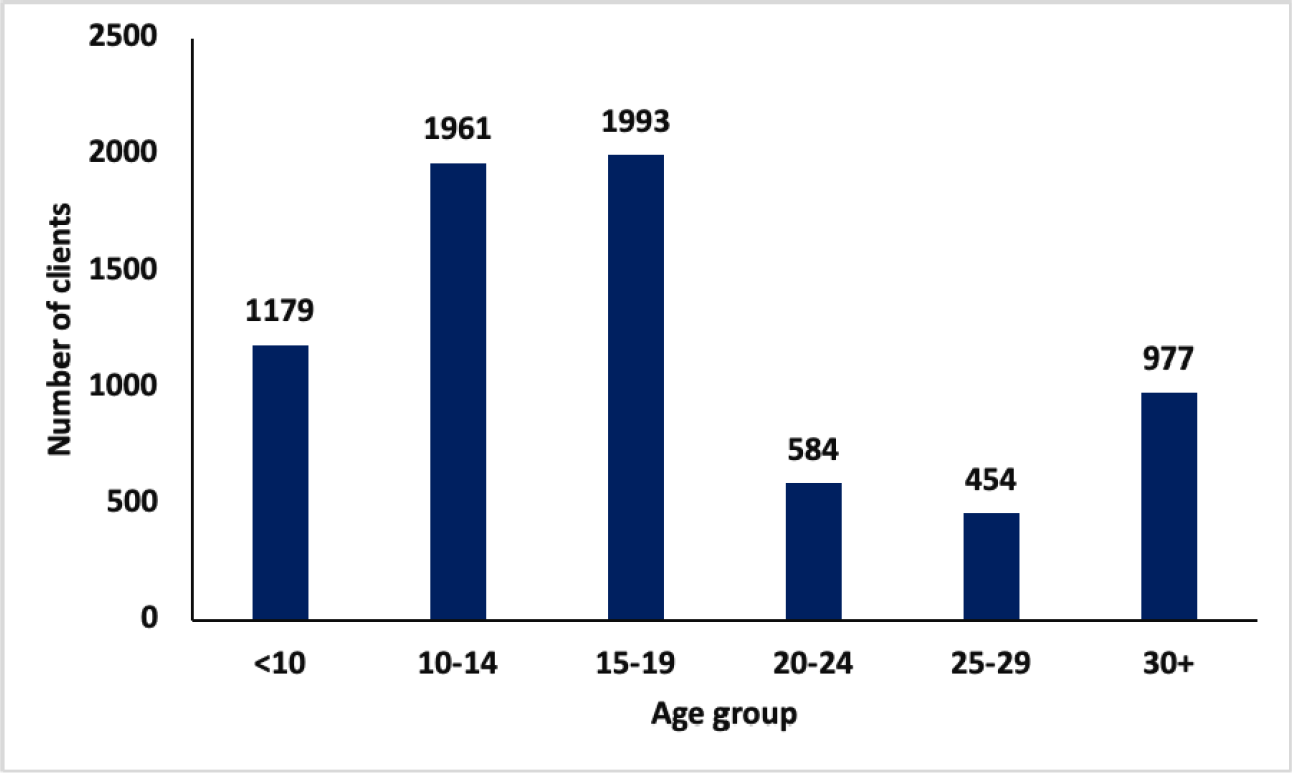
GBV cases by age group in all LH clinics from January 2020-September 2023.

## GBV by violence type

The type of violence reported includes physical, sexual physical, and emotional. In addition, a combination of the violence types can also be reported, such as physical and sexual, sexual and emotional, physical and emotional, and physical, sexual and emotional (PSE) violence. The most common form of violence reported was sexual violence, with 3,247 cases (45%), and the least common was combined physical and sexual violence, with 122 cases (22%) (see Figure 3). Emotional violence was the most frequently reported type of GBV for males (N=165), while sexual violence was the predominant form reported by females (N=3128) (see Figure 4).

**Figure 3:**
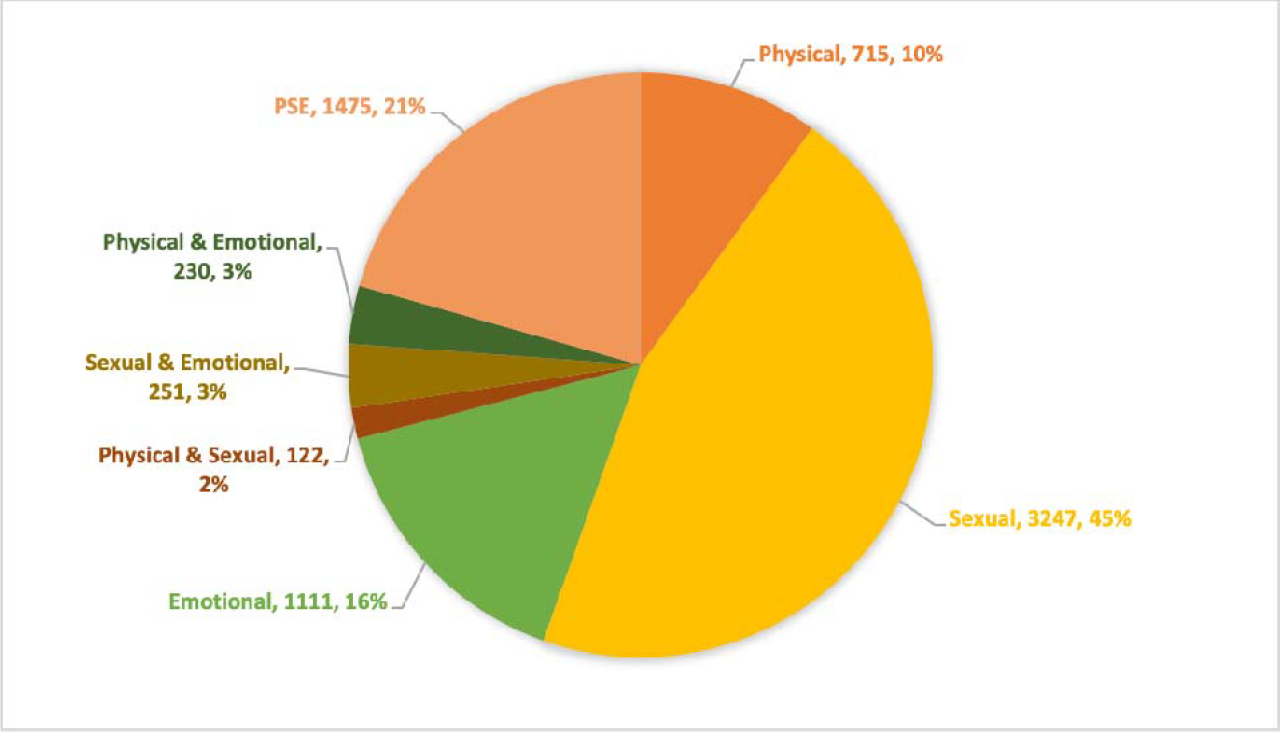
GBV by violence type.

**Figure 4:**
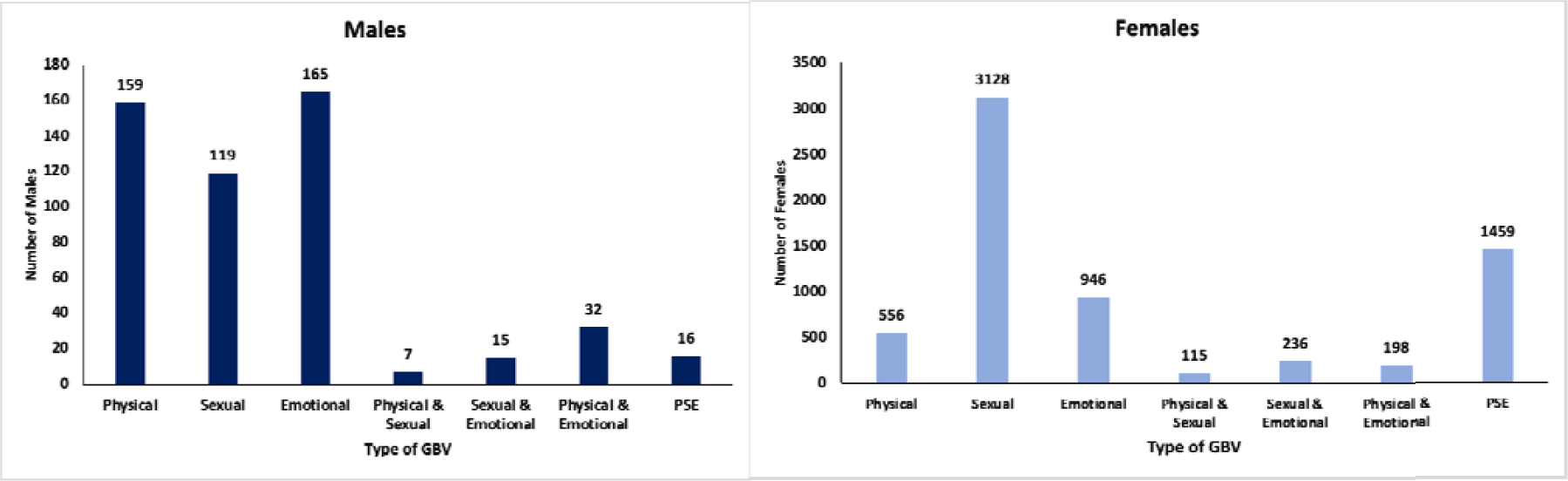
GBV by gender and violence type. PSE= physical, sexual, emotional, GBV=gender-based violence

## GBV trends

GBV cases increased from January 2020 to September 2023, especially among women and survivors aged 10-19 (see Figures 5 and 6). However, GBV cases among men have steadily been low.

**Figure 5:**
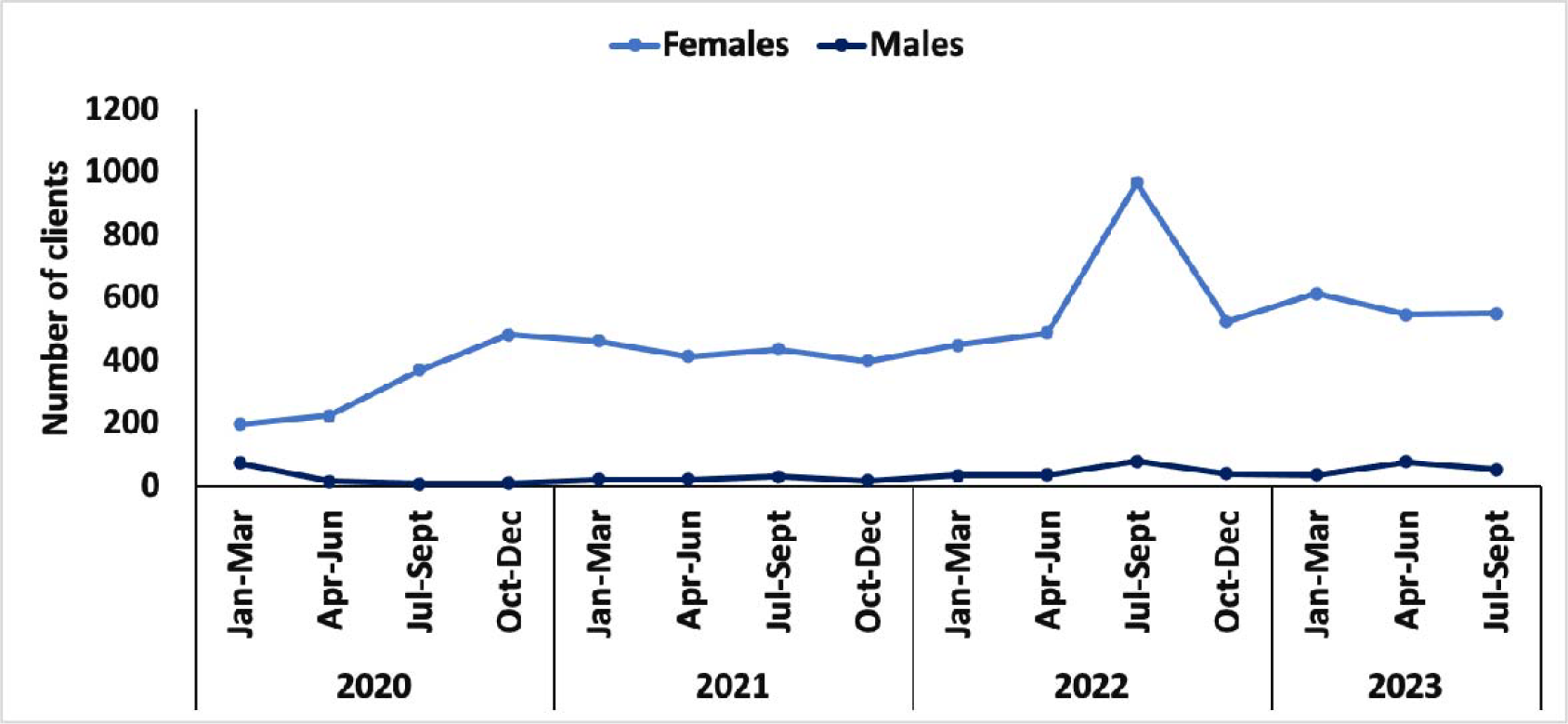
GBV trends by gender.

**Figure 6.**
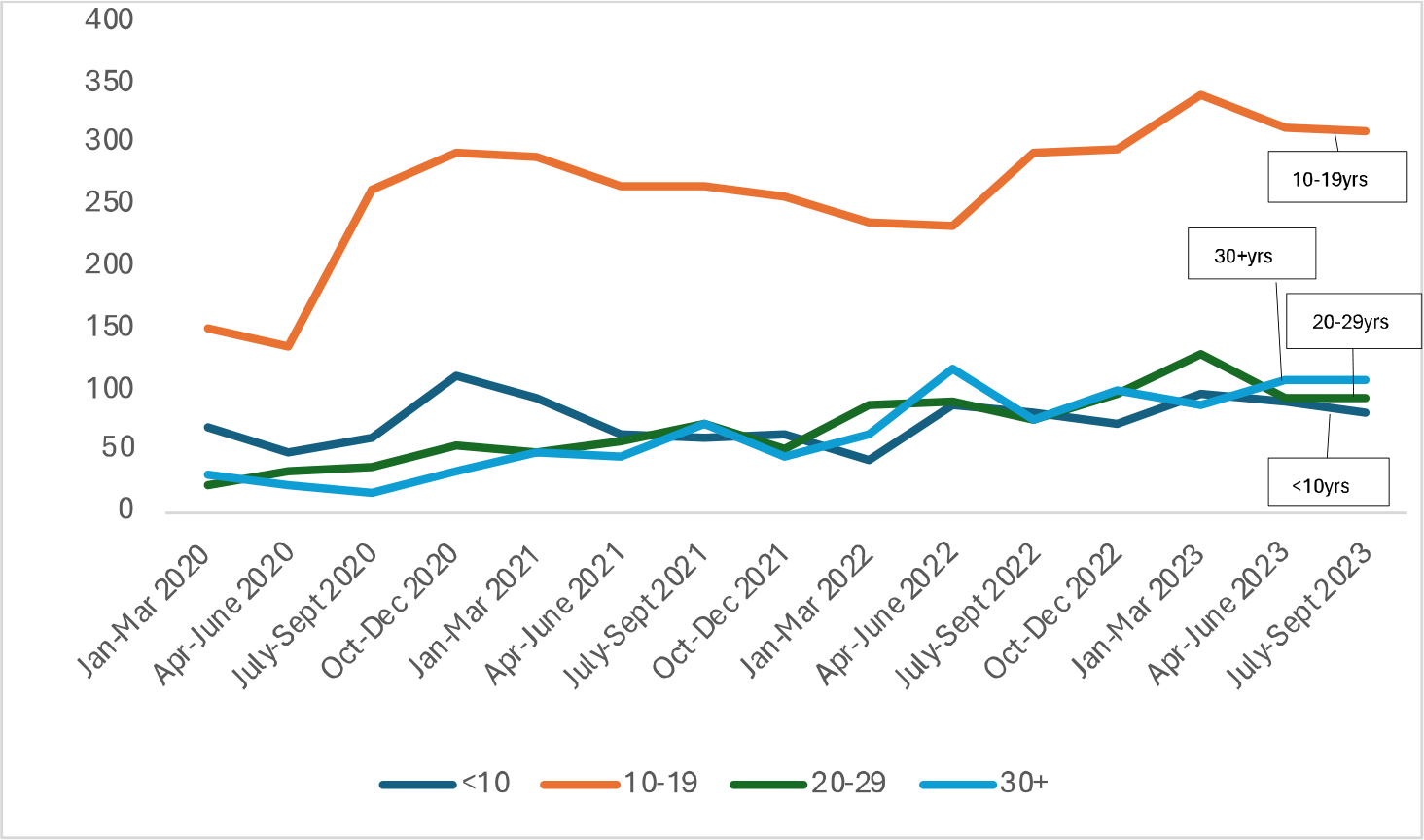
GBV trends by age group.

## Post-GBV care services

A total of 13,943 post-GBV services were documented. One survivor may receive more than one type of service depending on their needs. Overall, PSS was the most common type of service that was offered to GBV survivors (25%), followed by HTS (19%) and STI screening (18%) (see Figure 7).

**Fig. 7.**
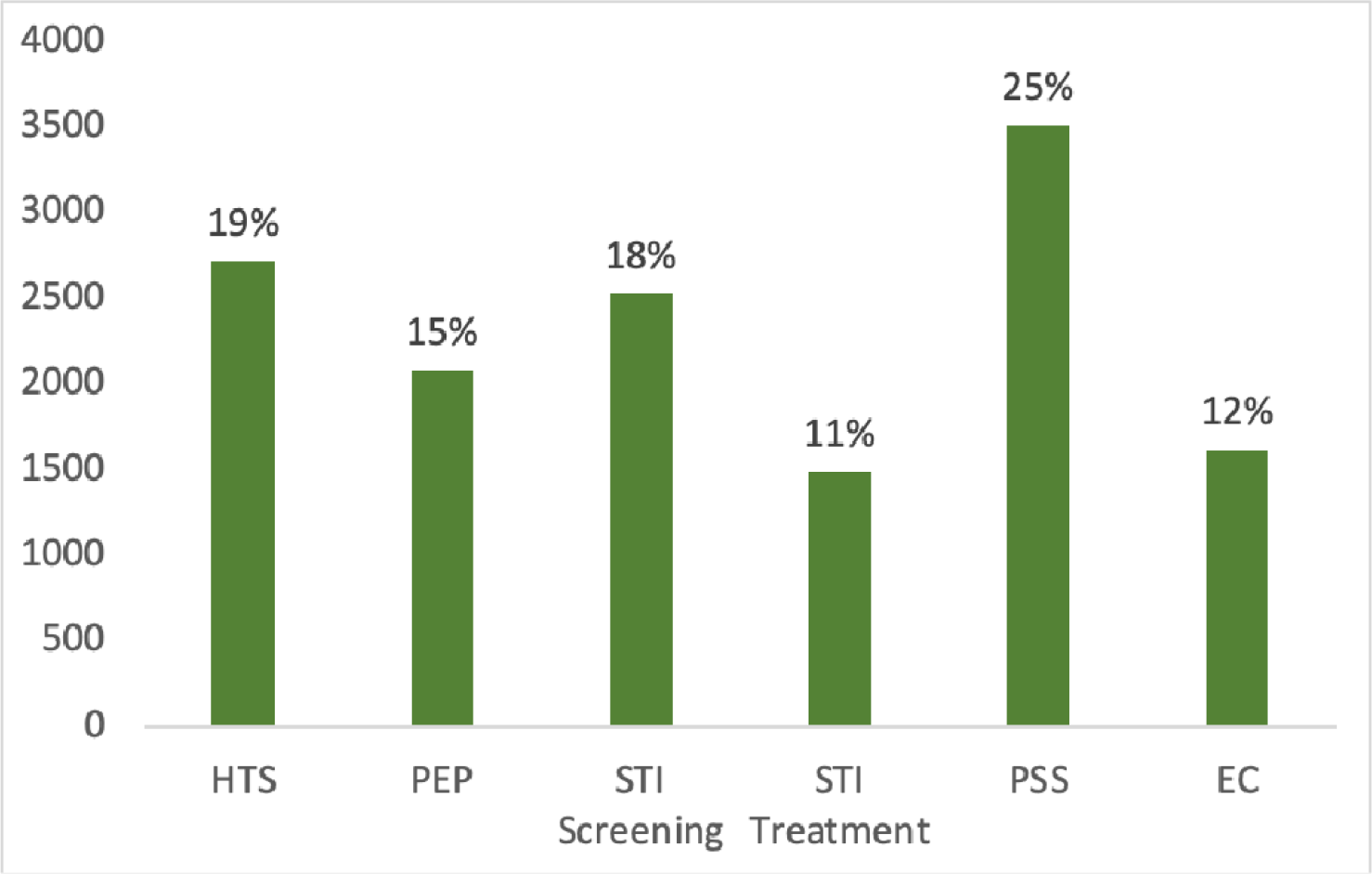
Post-GBV care services provided to survivors (N=13,943) HTS=HIV testing services, PEP=post-exposure prophylaxis, STI=sexually transmitted infections, PSS=psycho-social services, EC=emergency contraception

## Violence perpetrators

There were 3798 cases where the perpetrators of violence were reported as either known (parent, intimate partner, or other) or unknown to the survivor. The majority (44%) of the perpetrators of violence were other known assailants to the GBV survivor, followed by an intimate partner (34%), unknown assailants (12%), and parents (10%) (Table 1). Intimate partners constitute the most significant proportion of assailants physical violence (53%), combined physical and sexual (56%), and physical-emotional abuse cases (63%). The highest form of violence perpetrated by parents was emotional violence (30%). While the percentage of abuse by unknown assailants is relatively low, they still contribute notably to PSE (20%) and sexual violence (17%).

**Table 1:**
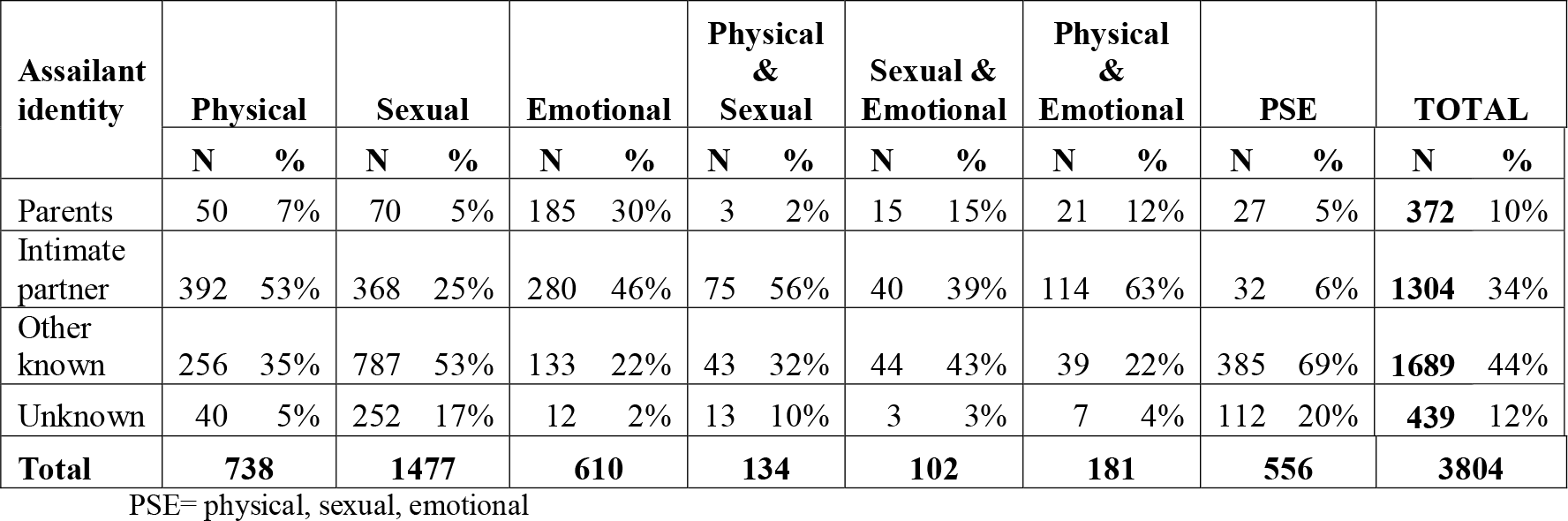
Assailant Identity by Type of Violence.

## Discussion

This paper describes integrating GBV services into routine ART clinic operations of Lighthouse Trust, Malawi’s largest ART provider. We outline how Lighthouse Trust effectively integrated GBV care into routine healthcare services by developing and implementing standardized procedures for screening, identification, care, referral, and documentation to optimally support survivors. These were further strengthened by strong collaboration with the OSCs. We noted a substantial number of reported cases, particularly affecting young women between 10 and 19 years old. This trend is in line with broader findings, such as those from the Violence Against Children and Young Women Survey, revealing widespread violence among this demographic, with sexual violence reported most frequently. We document a marked rise in GBV reports since July 2020, coinciding with the COVID-19 pandemic. The observed increase in GBV reporting aligns with global trends, exacerbated by the COVID-19 pandemic^27^., indicating not only the pervasiveness of the issue but also the critical need for such integrated care models^28^.

Consistent with global data^29^, our findings show a disproportionate number of female victims. However, this difference may not fully capture the prevalence of GBV, as studies suggest that men may be less likely to report such experiences due to factors like social stigma and gender norms^30,31^.

The majority of GBV survivors at Lighthouse Trust received psychosocial support alongside STI screening and HIV testing. Notably, most assailants were known to the victims but were neither parents nor intimate partners, with the latter constituting the second most frequent category of perpetrator. The high rates of known assailants is in concert with what others have reported^32–34^., and also highlights the critical need for secure refuge options for survivors, emphasizing the role of safe houses in GBV response frameworks^20^.

## Lessons Learned

Since 2001, Lighthouse Trust has integrated post-GBV care into its services, initially addressing only visible incidents or those disclosed by patients. To standardize this care, LH developed and implemented SOPs, training healthcare providers to screen and care for GBV survivors in a manner that would not retraumatize GBV survivors, echoing previous findings from similar settings^16,20^. This training also addressed a gap in providers’ recognition of emotional violence, leading to a broader understanding of its impact and acknowledgment of the importance of addressing all forms of GBV.

Even though all service providers were trained in GBV screening, the overwhelming workload led to inconsistent proactive screening by some. There were also gaps in making the necessary support service referrals. Cases were sometimes reported to the police prematurely, without the victims first receiving healthcare. Ideally, OSC would have on-site police officers to take reports. Still, staffing shortages mean victims often have to travel to police stations, discouraging some from reporting at all, leading to many unreported cases.

The register posed challenges in documenting multiple GBV types, such as physical, sexual, and emotional violence, leading to potential underreporting or misreporting. The focus was mainly on younger age groups, inadvertently omitting those over 30. Despite this, data on older survivors can be captured as ages are recorded. Additionally, heavy workloads led to incomplete documentation, suggesting that the true number of GBV cases may be higher than reported.

Instances arose where guardians hesitated to report sexual violence against child survivors to the police. In some cases, children were given money for sexual encounters with adults, mistakenly leading guardians to view the interactions as consensual, not considering the minors’ inability to consent legally. This highlights an urgent need for increased community awareness about GBV affecting young individuals^20^. Furthermore, the lack of a safe house posed a problem in cases where a child survivor was subject to abuse within their home, and no suitable relative could be found to provide a secure and protective environment. For example, one child was temporarily housed in a juvenile center for safety, awaiting a judicial decision for more permanent care. These scenarios emphasize the importance of providing secure shelters for minors who are survivors of GBV.

Some of the factors contributing to the increase in reported GBV cases include community sensitization efforts and GBV screening at ART clinics. Moreover, the collaborative efforts between Lighthouse clinics and the OSC have improved GBV case documentation. However, some clients may not receive the full post-GBV treatment package if they report to the facility beyond the recommended time, particularly those requiring PEP and emergency contraceptives.

LH maintains strong ties with the OSC, supporting them with HIV testing services and post-exposure prophylaxis. LH also supplements OSC staff with trained personnel when needed. During the pandemic, OSCs were temporarily converted into COVID-19 isolation centers, compelling GBV survivors to seek help in overcrowded hospitals, a situation that might have deterred some from pursuing services. Despite this, reports of GBV cases continued to increase.

## Recommendation

In conclusion, effective GBV care is vital for HIV patients, and training providers in LIVES enhances the providers’ abilities to identify and support GBV survivors. Increased awareness among providers and the community, particularly regarding minors, is crucial. Survivors should receive comprehensive GBV care, with efforts made to ensure service accessibility, confidentiality, and safety. Cross-departmental collaboration within health facilities is key for proper survivor support and referral.Furthermore, effective linkage, referral, and partnership between government agencies falling under different ministries, such as Health, Gender, Education, and Judicial, will help streamline GBV awareness, prevention, care, and support. This also includes the finding of resources to establish Safe Houses for GBV survivors who experienced violence in their homes, especially children.

Enhanced GBV services demand community engagement, strong connections with OSCs, and sustained psychological care for survivors. Our study underscores the urgency for Malawi’s policymakers to fortify protective measures and judicial processes, especially for minors, to ensure that GBV survivors receive safe, private, and comprehensive care. Implementing these strategies will strengthen GBV care in health facilities and foster a culture that upholds the safety and dignity of all patients, particularly GBV survivors.

## Data Availability

All data produced in the present study are available upon reasonable request to the authors.

## Acknowledgement

The authors would like to thank the One-Stop Centers and the Lighthouse GBV team for their unwavering commitment and contributions to this study. Their dedication to providing care for survivors has not only made this work possible but has also significantly advanced the support for those affected by gender-based violence.

